# Development of the Canadian Food Intake Screener to assess alignment of adults’ dietary intake with the 2019 Canada’s Food Guide healthy food choices recommendations

**DOI:** 10.1101/2022.12.19.22283569

**Authors:** Joy M. Hutchinson, Tabitha E. Williams, Ailish M. Westaway, Alexandra Bédard, Camille Pitre, Simone Lemieux, Kevin W. Dodd, Benoît Lamarche, Patricia M. Guenther, Jess Haines, Angela Wallace, Alicia Martin, Maria Laura da Costa Louzada, Mahsa Jessri, Dana Lee Olstad, Rachel Prowse, Janis Randall Simpson, Jennifer E. Vena, Sharon I. Kirkpatrick

## Abstract

The objective of this project was to develop a brief self-administered dietary screener, in English and French, to rapidly assess alignment of adults’ dietary intake with the 2019 Canada’ s Food Guide healthy food choices recommendations. In consultation with Health Canada and external advisors (n=15), foundational principles were defined. Existing screeners were scanned, and the healthy food choices recommendations were mapped to inform questions and response options. Cognitive interviews were conducted in English (n=17) and French (n=16) with adults aged 18-65 years from April to June 2021 to assess understanding of questions and face validity; recruitment emphasized variation in sociodemographic characteristics. Face and content validity were assessed with experts in nutrition, surveillance, and public health (n=13 English, 3 French) from April to May 2021. The testing indicated that the screener was well-understood overall but informed refinements to improve comprehension of the questions and their alignment with the healthy food choices recommendations. The resulting Canadian Food Intake Screener/Questionnaire court canadien sur les apports alimentaires includes 16 questions to rapidly assess alignment of intake with the 2019 Canada’ s Food Guide healthy food choices recommendations, including healthy foods and foods to limit, in situations in which comprehensive dietary assessment is not feasible.

**Novelty:**

- The Canadian Food Intake Screener was developed to rapidly assess alignment of adults’ dietary intake over the past month with the Food Guide’ s “healthy food choices” recommendations.
- The screener was developed and evaluated through an iterative process that included three rounds of cognitive interviews in each of English and French, along with ongoing feedback from external advisors and face and content validity testing with a separate panel of content experts.
- The 16-question screener is intended for use with adults, aged 18-65 years, with marginal and higher health literacy in research and surveillance contexts in which comprehensive dietary assessment is not possible.

## Introduction

Suboptimal dietary patterns are a key risk factor for noncommunicable chronic diseases in Canada and globally (Afshin et al. 2019; Lim et al. 2012; Lozano et al. 2012; Micha et al. 2017; Vajdi and Farhangi 2020). Diets low in whole grains, fruits, vegetables, nuts and seeds, and omega-3 fatty acids and high in sodium have been shown to be responsible for more deaths globally than any other risk factor (Afshin et al. 2019). To promote healthy eating and reduce diet-related chronic disease risk, many countries publish food-based dietary guidelines, presenting “specific, culturally appropriate, and actionable recommendations” (Food and Agriculture Organization 2022; Herforth et al. 2019). In 2019, Health Canada released an updated Canada’ s Food Guide (CFG-2019), with recommendations on “healthy food choices” and “healthy eating habits” (Health Canada 2021, Health Canada 2022a). In a shift from prior iterations, CFG-2019 does not provide recommendations on the number of servings per day or serving sizes for food groups based on age and sex. Instead, through the healthy food choices recommendations, CFG-2019 recommends eating a variety of healthy foods each day, including fruits and vegetables, whole-grain foods, and protein foods, emphasizing more frequent consumption of plant-based protein foods. The CFG-2019 plate provides a visualization of the desirable proportions of foods from these categories in relation to one another (Health Canada 2022a). CFG-2019 aims to promote healthy eating and overall nutritional well-being and to support improvements to the food environment in Canada (Health Canada 2022b).

For research and surveillance purposes, it is of interest to assess alignment of eating patterns and practices with CFG-2019, for example, to inform targeted interventions to address disparities in alignment with the guidance among population subgroups. The Healthy Eating Food Index-2019 (HEFI-2019) facilitates assessment of the alignment of dietary intake with CFG-2019 healthy food choices recommendations in research and surveillance contexts in which comprehensive dietary intake data, such as from 24-hour dietary recalls, are available (Brassard et al. 2022a, 2022b). Data from dietary recalls are recommended for characterizing the dietary intake of populations and subgroups due to their comprehensiveness, as well as their greater accuracy relative to frequency-based instruments (Freedman et al. 2014; Freedman et al. 2015; Kirkpatrick et al. 2022a; National Cancer Institute 2015; Thompson et al. 2015). However, while online self-administered recall platforms have eased researcher and respondent burden (Lafrenière et al. 2017; Subar et al. 2012), recalls can be time consuming to collect and may not be amenable to all settings. Further, appropriate use of recall data requires substantial expertise and extensive cleaning and analytic efforts (Kirkpatrick et al. 2022b).

In contrast, brief dietary questionnaires, informally called “screeners,” can be used for rapid assessment of food and beverage intake over a given period, such as the past month or year (National Cancer Institute 2015, Thompson et al. 2015). Screeners often focus on specific dietary components (e.g., fruits and vegetables, fibre) (Centers for Disease Control and Prevention n.d.; Hedrick et al. 2010; Tangney et al. 2019) but may be multifactorial (Colby et al. 2020; de Rijk et al. 2021; Fulkerson et al. 2012; Gnagnarella et al. 2018; Lafrenière et al. 2019; Thompson et al. 2004). Prior multi-factorial screeners have aimed to assess dietary intake relative to food-based dietary guidance (Colby et al. 2020; de Rijk et al. 2021; Gabe and Jaime 2019).

The objective of this study was to develop a brief screener to assess overall alignment of adults’ dietary intake with CFG-2019 healthy food choices recommendations. The screener was developed for use with adults, aged 18-65 years, with marginal and higher health literacy, and is intended for self-administration in English and French. The current paper describes the development process, including cognitive testing to assess whether screener questions were understood as intended, and face and content validity with a panel of experts. The screener’ s scoring system and construct validity are described in the accompanying paper (Hutchinson et al. 2022). A separate brief questionnaire, the Canadian Eating Practices Screener, developed to assess adults’ alignment with the CFG-2019 healthy eating habits recommendations, is described elsewhere (Haines et al. 2022).

## Materials and Methods

### Development of the screener

Screener development and evaluation were undertaken in collaboration with Health Canada and guided by a team of external expert advisors, including nutrition researchers and practitioners (**Supplementary File S1**); this group included English- and French-speaking individuals. Many of the advisors were involved in the development of the HEFI-2019 (Brassard et al. 2022a, 2022b), supporting consistent interpretation of the underlying dietary guidance and alignment of the screener with the HEFI-2019.

The development of the screener drew upon the messaging related to healthy food choices in CFG-2019, including the plate depicting the recommended proportions of the food categories (Health Canada 2022a). Also considered were the food choice components within the Healthy Eating Recommendations, which provide simple and actionable messages for consumers (Health Canada 2020), and the Dietary Guidelines, which are intended primarily for health professionals and policymakers (Health Canada 2022b). In addition to the guidance, the components included in the HEFI-2019 and their construction (e.g., inclusion or exclusion of particular foods) (Brassard et al. 2022a, 2022b) informed the screener questions to some extent. We also drew upon available information on the dietary intake of Canadians (e.g., key sources of food categories and nutrients such as saturated fats) (Harrison et al. 2019; Kirkpatrick et al. 2019a; Tugault-Lafleur and Black 2019).

The steps in screener development are illustrated in **Figure 1** and described below, though the process was iterative in terms of consultation with the advisors and refinement of the screener.

**Figure 1.**
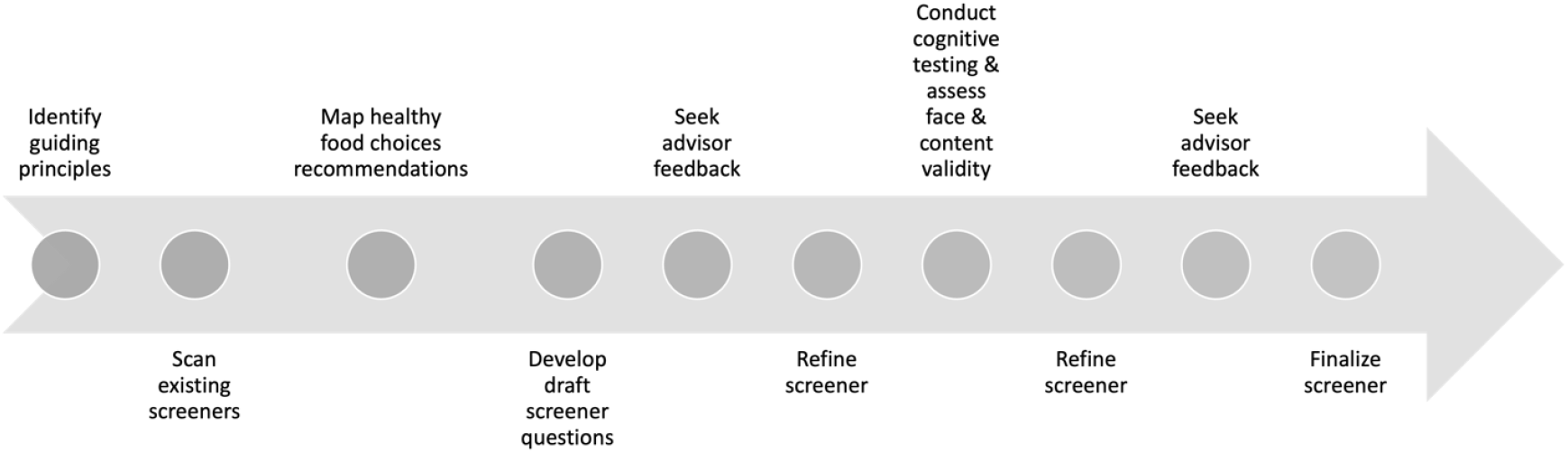
Process for development of the Canadian Food Intake Screener to assess alignment of adults’ intake with the healthy food choices recommendations.

#### Defining guiding principles for the screener

The development of the screener was informed by guiding principles, defined in collaboration with Health Canada *a priori* (**Box 1**). These principles related to the development of a brief screener that is simple to use and score, assesses alignment with the healthy food choices recommendations overall, and considers the numeracy and literacy levels of the target population. Equivalence or comparability (Frongillo et al. 2019) in capturing alignment with the CFG-2019 healthy food choices recommendations across population subgroups was also considered. Additionally, the screener should demonstrate reasonable construct validity, described in the accompanying paper (Hutchinson et al. 2022). These principles were discussed with the advisors and informed initial decisions about the format and content of the screener.

##### Box 1.

Guiding principles for the development and evaluation of the Canadian Food Intake Screener

- Simple to use and score.
- Brief (<10 minutes).
- Assess adherence to the 2019 Canada’ s Food Guide healthy food choices recommendations overall, not specific recommendations.
- Consider the numeracy and literacy levels of the target population.
- Consider equivalence (i.e., comparability) in capturing the construct across subgroups of the target population.
- Demonstrate reasonable validity for capturing the construct in the target population.

These decisions included assessing frequency of consumption versus proportions, capturing foods and beverages versus nutrients, and assessment of alignment with the healthy food choices recommendations overall.

First, although the CFG-2019 plate identifies the recommended proportions to be contributed by vegetables and fruit, whole-grain foods, and protein foods, the screener does not focus on proportionality. This is because screeners do not capture total dietary intake so a denominator, which would be needed to calculate proportions allocated to different types of foods, is unavailable. To capture usual consumption, participants could be asked to average proportions across eating occasions over some period, such as a month. However, this approach was hypothesized to be cognitively challenging, as well as difficult to score.

Alternatively, repeat administrations of a screener focused on proportions on a given day could be used to capture usual proportions, but this would add burden for researchers and participants, undermining the goal of a brief screener. Furthermore, not all CFG-2019 recommendations related to healthy food choices are expressed using proportionality, such as the recommendations to limit intake of highly processed foods. Given the guiding principles related to literacy and numeracy demands, a frequency-based screener was thus developed, with the hypothesis that patterns of frequency of intake of different foods and beverages would provide an indication of the degree of alignment with the CFG-2019 healthy food choices recommendations. The past month, which is the typical period queried by screeners (Centers for Disease Control and Prevention n.d.; England et al. 2017; National Cancer Institute 2021; Wijnhoven et al. 2018), was selected as the time frame of interest. A focus on the past month provides an indication of longer-term intake (versus intake on a given day) and may reduce error compared to recalling and averaging frequency of consumption over a longer period, such as a year (National Cancer Institute 2015). The screener does not query portion sizes, which is common for brief instruments (National Cancer Institute 2015).

Second, the screener focuses on frequency of intake of foods and beverages and not nutrients. The guidance is primarily food-based, though the Dietary Guidelines include recommendations related to intake of free sugars, saturated fats, and sodium (Health Canada 2022b). A screener specifically focused on one of these nutrients would likely include many questions and even then, may not accurately estimate nutrient intake (Tangney et al. 2019). Per the guidance and examinations of dietary intake among the population (Harrison et al. 2019; Kirkpatrick et al. 2019a), highly processed foods account for high proportions of intake of sugars, saturated fats, and sodium; thus, questions on highly processed foods were expected to provide a moderately strong signal in terms of the extent of alignment of dietary intake with the healthy food choices recommendations.

Finally, it was determined that scoring should focus on alignment with the recommendations overall, given that a brief multi-factorial screener cannot, by design, provide accurate estimates of intake of particular food categories.

#### Scanning existing screeners, mapping the healthy food choices recommendations, developing screener questions, and soliciting feedback from advisors

Screener development and evaluation were informed by existing screeners that have undergone validation (Centers for Disease Control and Prevention n.d.; Colby et al. 2020; England et al. 2017; Gadowski et al. 2020; Gnagnarella et al. 2018; National Cancer Institute 2019; Tangney et al. 2019). These screeners provided insights into format and possible questions for inclusion, as well as response options. CFG-2019 recommendations pertaining to healthy food choices were then mapped (**Table 1**) to provide a roadmap against which to develop screener questions to ensure content validity. Initial screener questions were developed and revised iteratively based on feedback from Health Canada and the advisors. The screener was developed in English and translated to French. Translations were conducted by a professional firm and reviewed by bilingual researchers at Université Laval.

**Table 1:** Dietary guidance mapped to final questions within the Canadian Food Intake Screener, in English, to assess alignment of intake with the 2019 Canada’ s Food Guide healthy food choices recommendations among adults aged 18-65 years (NOTE: questions in the table are not in order, refer to Supplementary File S3 for the final screeners in English) ^1^.

| Dietary Guideline | Healthy Eating Recommendation and/or other guidance | Final English screener questions |
| --- | --- | --- |
| Vegetables, fruit, whole grains, and protein foods should be consumed regularly | Eat plenty of vegetables and fruit | Over the past month, how often did you eat potatoes, including baked, boiled, or mashed potatoes, or sweet potatoes? **Do not include** french fries, poutine, home fries, or hash browns. |
|  |  | Over the past month, how often did you eat fresh, cooked, frozen, or canned vegetables? **Do not include** potatoes, french fries, poutine, or other deep-fried vegetables, or vegetable juices and drinks. |
|  |  | Over the past month, how often did you eat fresh, frozen, canned, or dried fruit? **Do not include** fruit juices and drinks. |
|  | Eat whole grain foods | Over the past month, how often did you eat **whole wheat or whole grain** breads, bagels, pasta, noodles, quinoa, oats, brown or wild rice, breakfast cereals, or other whole wheat or whole grain foods? **Do not include** white breads, bagels, pasta, noodles, rice, or refined breakfast cereals. |
|  | Eat protein foods | Over the past month, how often did you eat eggs, beef, pork, wild meat, chicken or other poultry, fish, shellfish, or other animal-based sources of protein? Include canned fish and canned poultry. **Do not include** fast food, hot dogs, sausages, beef jerky, bacon, ham, or other deli or luncheon meats. |
|  |  | Over the past month, how often did you eat nuts, seeds, tofu, beans and lentils, peanut butter or other nut butters, or other plant-based sources of protein? **Do not include** green beans or packaged veggie burgers and plant-based meats. |
|  |  | Over the past month, how often did you eat yogurt, kefir, or cheese? |
| Among protein foods, consume plant-based more often | Choose protein foods that come from plants more often | Over the past month, how often did you eat nuts, seeds, tofu, beans and lentils, peanut butter or other nut butters, or other plant-based sources of protein? **Do not include** green beans or packaged veggie burgers and plant-based meats. |
| Water should be the beverage of choice | Make water your drink of choice<br>Replace sugary drinks with water | The screener does not assess water consumption. |
|  | Healthy drink options other than water can include:<br>White milk (0% and 1% milk) | Over the past month, how often did you have **white** cows' milk or **unsweetened** plant-based beverages (e.g., soy, almond, or oat milk)? **Do not include** small amounts in coffee or tea, or chocolate and other sweetened milk. |
|  | Unsweetened fortified plant-based beverages such as soy beverage or almond beverage<br>Unsweetened coffee and tea | Over the past month, how often did you have chocolate milk or other **flavoured** milk or **sweetened** plant-based beverages (e.g., soy, almond, or oat milk)? **Do not include** small amounts in coffee or tea, or diet/artificially sweetened or sugar-free beverages. |
|  |  | Over the past month, how often did you drink fruit juice, fruit-flavoured drinks, soda or pop, **sweetened** sports or energy drinks, **sweetened** hot or iced coffee or tea, or **sweetened** waters? **Do not include** diet/artificially sweetened or sugar-free beverages, such as diet soda. |
| Foods that contain mostly unsaturated fat should replace foods that contain mostly saturated fat | Choose foods with healthy fats instead of saturated fats | Over the past month, how often did you have margarine or vegetable oils (e.g., olive, canola, or sunflower oil)? **Do not include** lard, coconut oil, palm oil, or butter. |
| Processed or prepared foods and beverages that contribute to excess sodium, free sugars, or saturated fat undermine | Limit highly processed foods. If you choose these foods, eat them less often and in small amounts.<br>Prepare meals and snacks using | Over the past month, how often did you eat hot dogs, sausages, beef jerky, bacon, ham or other deli or luncheon meats? **Do not include** fast food, canned fish, canned poultry, or packaged veggie burgers and plant-based meats. |
| healthy eating and should not be consumed regularly | ingredients that have little to no added sodium, sugars or saturated fat<br>Choose healthier menu options when eating out | Over the past month, how often did you eat food from fast food restaurants, such as burgers, french fries, poutine, pizza, submarine sandwiches, fried chicken, burritos, or tacos? |
|  |  | Over the past month, how often did you eat cookies, cakes, muffins, pastries, granola bars, protein bars, ice cream, candy, chocolate, sugary breakfast cereals, or other sugary foods? |
|  |  | Over the past month, how often did you eat crackers, chips, pretzels, popcorn, or other salty snacks? |
|  |  | Over the past month, how often did you eat **white** breads, bagels, rice, pasta, noodles, or other refined grains, such as breakfast cereals? **Do not include** whole wheat or whole grain foods. |

<sup>1</sup>The full version of this table, including French, is not accessible here due to server language requirements. The full table is available from the corresponding author upon request.

Given the guiding principles, a key emphasis in seeking advisor feedback on the draft questions was weighing trade-offs between a nuanced screener mapped closely to the recommendations versus a simple screener. It was expected that more detail would result in higher cognitive load and accordingly, more reporting error (Natarajan et al. 2010). This reporting error may be differential between individuals with different characteristics, for example, with respect to literacy (Choi and Cawley 2018; Keogh et al. 2020), resulting in less utility of the screener for use with diverse populations and potentially masking differences in alignment with the guidance among population subgroups. There was accordingly consensus among the advisors that the screener did not need to capture every nuance of the guidance (e.g., differentiating fruits canned in syrup from those not canned in syrup, capturing specific sources of unsaturated fats such as avocado); such nuances can be more adequately addressed using more comprehensive methods, such as 24-hour dietary recalls.

The iterative feedback from the advisors led to consensus on wording questions as simply as possible, avoiding technical terms (e.g., fortified); querying foods of interest using colloquial terms (e.g., plant-based milks); including examples of commonly consumed foods and relevant exclusions but avoiding lengthy, exhaustive lists; using consistent question structure and response options; and ordering questions such that earlier questions cue responses to later questions. We sought to avoid combining different types of foods (e.g., meats, cheese, and milk) in a single question to the extent possible, while also aiming for a short screener. This approach was deemed useful for minimizing cognitive load and ensuring clarity in the foods to consider in responding to each question, as well as providing flexibility to account for emphases of the guidance, for example, on plant-versus animal-based protein foods, in the screener’ s scoring system, described in the accompanying paper (Hutchinson et al. 2022). The specific wording used in the screener questions differs in some instances from language used in the guidance and the use of wording such as ‘sources of’ does not imply that the foods or beverages must meet the nutrient criteria for the claim “source of”, as defined in the Food and Drug Regulations (Government of Canada 2022).

The version of the screener evaluated in the first round of cognitive testing and in face and content validity testing consisted of 15 questions (**Supplementary File S2**). Response options were adapted from the Dietary Screener Questionnaire and the Diet History Questionnaire (National Cancer Institute 2021; National Cancer Institute 2022; Millen et al. 2006; Subar et al. 2001; Thompson et al. 2017) and ranged from never to 6 or more times per day.

#### Cognitive testing

Cognitive testing is a qualitative, psychologically oriented method to investigate the ways in which research participants interpret and respond to survey questions, typically via individual interviews (Foddy 1996; Willis 2005; Willis and Artino 2013; Willis and Miller 2011). The goal is to determine whether each question is understood consistently in the way researchers intend (Collins 2003; Beatty and Willis 2007; Willis and Artino 2013). Cognitive testing draws upon insights from psychology regarding the cognitive processes involved in responding to survey items. Tourangeau’ s four-stage model describes these processes, which include 1) comprehension, 2) retrieval of information, 3) judgment or estimation, and 4) selection of a response to the question (National Research Council 1984). Most cognitive testing procedures target the level of comprehension because this is where problems most often occur (Foddy 1996; Willis et al. 2013). A cognitive interviewing reporting framework proposed by Boeije and Willis (2013) was used to guide reporting of this aspect of the process.

#### Data collection

Cognitive interviews were conducted from April to June 2021. The interviews in English were conducted by researchers at the University of Waterloo and those in French by researchers at Université Laval. Interviews in each language were led by researchers (TEW and AB) with training in qualitative methods. Ethics review and approval was obtained from the University of Waterloo Office of Research Ethics (ORE #42994), the Université Laval Research Ethics Board (REB #2021-088), and the Health Canada and Public Health Agency of Canada Research Ethics Board (REB #2020-044H).

While cognitive testing is often conducted informally, research suggests small sample sizes may fail to detect problems with survey questions, including those that may introduce measurement error (Blair et al. 2006). Recommendations for sample size range from 10 to 30 total participants, or five to 15 participants per round for two to three rounds (Beatty and Willis 2007; Willis and Artino 2013; Willis and Miller 2011). To adequately probe for potential issues with comprehension of the screener with individuals with varied sociodemographic characteristics, a sample of approximately 32 participants, to take part in 16 interviews in English and 16 in French, was sought. Interviews were conducted in three rounds in each language, with four to eight participants per round, to allow for iterative refinement of the screener (Beatty and Willis 2007).

Potential participants were recruited through community organizations and social media (English), and through community organizations and a database of potential research participants (French). For the testing in English, potential participants completed an eligibility questionnaire hosted on Qualtrics (Qualtrics, Provo, UT). For the testing in French, potential participants completed an eligibility questionnaire and returned it to the research coordinator via email. Eligible individuals were aged 18-65 years, lived in Canada, and were able to read the screener and complete a 45–60-minute interview using online teleconferencing software in English or French. The eligibility questionnaire also captured information on age, gender identity, racial/ethnic identity, educational attainment, and perceived income adequacy. Quota sampling was used to seek a balance of participants with varying educational attainment.

Specifically, the aim was for half of participants in the study to have not completed post-secondary education, as a proxy for lower literacy levels (Laramee et al., 2007; Schillinger et al., 2006). An approximate balance between women and men was sought, with a desire to include some individuals identifying as non-binary. Purposive sampling was used to maximize variation in other sociodemographic characteristics, including age, racial/ethnic identity (Black, East/Southeast Asian, Indigenous, Latino, Middle Eastern, South Asian, White), and perceived income adequacy (“Thinking about your total monthly income, how difficult is it for you to make ends meet?”, with response options including very difficult, difficult, neither easy nor difficult, easy, and very easy) (Litwin and Sapir 2009).

Eligibility questionnaire data were reviewed relative to the quotas and purposive sampling criteria on an ongoing basis, and eligible individuals were invited by email to participate in an interview. Those who agreed were sent an information letter and an informed consent form in advance. These documents were reviewed at the beginning of the interview, at which time the participant was asked to provide verbal informed consent. Interviews in English were conducted using Zoom (Zoom Video Communications, San Jose, CA), except for one conducted by telephone because the participant did not have Internet access, and interviews in French were conducted using Microsoft Teams (Microsoft Corporation, Redmond, WA). A note-taker captured details of participants’ responses during the interview. Most interviews were audio-recorded, with participant consent, to allow researchers to review recordings as needed. Participants who completed an interview received a $20 CAD honorarium via Interac e-transfer in appreciation of their time. Following each interview, the interviewer and note-taker completed a debriefing form to note overall impressions and reflections on the interview process. By the end of the third round in each language, diminishing returns (Beatty and Willis 2007) were noted, in that few new problems were being identified, and recruitment and data collection were concluded.

#### Cognitive interview guide

Because the screener is intended for self-administration, participants were asked to complete it independently before they reviewed each question and their response process with the interviewer using a think-aloud approach (e.g., “can you walk me through how you arrived at that number?”) (Beatty and Willis 2007; Lenzner et al. 2016). The interviewer then used open-ended verbal probes to gauge understanding and thought processes, using a semi-structured interview guide developed in English and translated to French. The probes were aligned with the cognitive stages of processing. For example, processing of the question, “In the past month, how often did you consume fresh, frozen, and canned fruit?”, requires the respondent to understand and interpret keywords and phrases, including “how often”, “consume”, and “fruit”; to recall the correct response by thinking about how frequently they consumed fruit in the past month and to make a judgment about what number to report; and finally, to provide a response that matches the options available within the screener (e.g., “2 times per week”). The corresponding probes asked, “are there any words or ideas in this question that were difficult to understand?”, targeting comprehension, and “how sure or unsure are you that the number you provided is accurate?”, targeting retrieval of information and judgment. Sorting of foods and beverages across questions and assessment of face and content validity were integrated by asking respondents what kinds of fruits they thought of and those they excluded when answering each question. This process was repeated for each screener question.

To ensure comparability of testing approaches (Willis and Miller 2011), the English- and French-speaking teams used consistent interview guides, and the lead interviewer from the French-speaking team observed pilot interviews conducted in English with graduate students not involved in this research.

#### Data analysis

As is common in cognitive testing (Willis 2005; Beatty and Willis 2007; Willis and Artino 2013; Willis and Miller 2011), the interview notes and recordings were informally coded. After each round of interviews in each language, issues that may have required changes to ensure that questions were understood as intended were identified and summarized (Lenzner et al, 2016; Willis and Miller 2011). The two teams debriefed between rounds and following the final round and modified the screener iteratively to address issues identified in each language. To ensure translational equivalency, a decentering approach that recognized that problems identified in one language may require changes to the screener in both languages was applied (Brislin 1970; Willis et al. 2008). Prior to the final round of testing in English and the second round of testing in French, issues that had arisen were discussed with the advisors, who provided feedback and suggested modifications to the screener for the next round of interviews.

For reporting purposes, the issues identified were subsequently grouped into problem categories, or themes related to cognitive processes (Bobrovitz et al, 2015; Lenzner et al, 2016; Thompson et al, 2022; Willis and Miller 2011).

#### Face and content validity testing

Face and content validity testing was conducted to examine whether the screener was well-constructed and grounded in an understanding of the underlying phenomenon of interest (Frongillo, et al. 2019; Kirkpatrick et al. 2019b). This testing was completed by Health Canada from April to May 2021. Experts in nutrition, surveillance, and public health were identified by Health Canada based on existing contacts, including individuals who participated in the process to update the CFG-2019 and/or other relevant projects, and invited by email to participate.

Advisors involved in the development of the screener (Supplementary File S1) were not invited to participate in this phase. Ethics approval was obtained from the Health Canada and Public Health Agency of Canada Research Ethics Board (REB# 2020-044H). According to the associated policies for ethical research conduct, the face and content validity experts were not considered participants since they were not themselves the focus of the research; therefore, informed consent was not required.

Content experts who agreed to participate were sent, via email, the version of the screener tested in the first round of cognitive testing (Supplementary File S2) and a Microsoft Excel (Microsoft Corporation, Redmond, WA) spreadsheet prompting them to comment on whether each screener question reflected the guidance it was intended to capture and was easy to understand, with space for comments. The spreadsheet was returned to Health Canada staff via email and the results summarized, including identifying questions that did not perform well according to multiple experts, as well as any global feedback on the screener. These results were shared with the cognitive testing teams and informed modifications to the screener in advance of the final rounds of cognitive interviews in each language.

## Results

### Cognitive testing

For the interviews in English, 193 potential participants completed the eligibility questionnaire, of whom 136 met the eligibility criteria. A total of 22 individuals were contacted, and 17 completed an interview. For recruitment of French-speaking participants, 101 potential participants completed the eligibility questionnaire, of whom 97 were eligible, and 16 were contacted and completed an interview. A total of 33 interviews (17 in English and 16 in French) were conducted. In total, 21 participants identified as women and 12 identified as men (**Table 2**). No participants identified their gender as non-binary. Participants represented a mix of racial identities, though the majority (n=19) identified as White. About half of the participants (n=13) had less than post-secondary education.

**Table 2:** Characteristics of participants (n=33) in cognitive interviews in English and French to evaluate the comprehension and face validity of the Canadian Food Intake Screener.

| Characteristic | English | French | Total |
| --- | --- | --- | --- |
|  | n |  |  |
| **Total** | 17 | 16 | 33 |
| **Age (years)** |  |  |  |
| 18-24 | 3 | 3 | 6 |
| 25-34 | 5 | 3 | 8 |
| 35-44 | 2 | 3 | 5 |
| 45-54 | 3 | 3 | 6 |
| 55-65 | 4 | 4 | 8 |
| **Gender identity** |  |  |  |
| Man | 5 | 7 | 12 |
| Woman | 12 | 9 | 21 |
| **Racial identity** |  |  |  |
| White | 9 | 10 | 19 |
| Indigenous | 0 | 4 | 4 |
| East/Southeast Asian | 4 | 0 | 4 |
| Black | 2 | 0 | 2 |
| Middle Eastern | 0 | 1 | 1 |
| South Asian | 1 | 0 | 1 |
| Latino | 0 | 1 | 1 |
| Prefer not to answer | 1 | 0 | 1 |
| **Perceived income adequacy<sup>1</sup>** |  |  |  |
| Very easy | 2 | 2 | 4 |
| Easy | 5 | 4 | 9 |
| Neither easy nor difficult | 6 | 8 | 14 |
| Difficult | 3 | 2 | 5 |
| Very difficult | 1 | 0 | 1 |
| **Educational attainment<sup>2</sup>** |  |  |  |
| High school graduate | 2 | 7 | 9 |
| Some college | 1 | 0 | 1 |
| Some university | 4 | 0 | 4 |
| College graduate | 2 | 3 | 5 |
| University graduate | 6 | 6 | 12 |
| Postgraduate training or degree | 2 | 0 | 2 |

<sup>1</sup> Perceived income adequacy was based on a question that asked, “Thinking about your total monthly income, how difficult is it for you to make ends meet?” (Litwin and Sapir 2009).
<sup>2</sup>The French eligibility questionnaire did not include “Some college”, “Some university”, or “Postgraduate training or degree” as response options.

Issues with the screener questions identified during cognitive testing generally fell into one of four themes, mainly related to comprehension, described below.

#### Lack of clarity about what to include or exclude

Lack of clarity about what to include or exclude in responses to screener questions related to groupings of foods and forms of foods that can be consumed in different ways. With respect to food groupings, in some cases, participants were uncertain about the types of foods that should be reported in response to a given question. Such problems most often occurred due to a lack of detail or examples in a question (**Table 3, example 1**). Contrarily, confusion also occurred when examples were too specific, as participants were unsure whether to “think outside the box” or report only the foods listed (Table 3, **examples 2 and 3**). These ambiguities were addressed by adding detail to existing questions or adding new questions to the screener, as well as through formatting and ordering of questions.

**Table 3.** Examples of issues identified in cognitive interviews in English and French to evaluate the comprehension and face validity of the Canadian Food Intake Screener^1^.

| Example | Problem category | Focus of question | Cognitive issue | Modifications |
| --- | --- | --- | --- | --- |
| 1 | Lack of clarity about what to include or exclude/Food groupings | Vegetables | Several participants did not consider potatoes in their response related to frequency of consuming vegetables. Participants explained they view potatoes as a starch rather than a fresh vegetable, and some felt potatoes should be reported with grain foods instead. | A question probing potato consumption, prior to the vegetable question, was added.<br>This modification was tested in the final rounds of cognitive testing in English and French and appeared to help participants understand where to include potato consumption. |
| 2 | Lack of clarity about what to include or exclude/Food groupings | Pre-made and ready-to-eat meals | The initial version of the screener asked, “How often did you consume deep-fried foods and ready-to-heat or ready-to-eat dishes?” (in French, “Au cours du dernier mois, à quelle fréquence avez-vous consommé des aliments frits et des plats prêts à réchauffer ou prêts à manger?”).<br>Several participants expressed difficulty understanding which foods to include in their responses to this question. | “Ready-to-heat or ready-to-eat” was simplified to “pre-made meals”. However, in the next rounds of interviews, participants were uncertain whether all take-out foods, or only deep-fried and fast foods, should be reported.<br>This question was then replaced with a more specific one probing frequency of consumption of fast foods, such as pizza, burgers, and French fries, to clarify inclusion criteria, while aligning with examples of highly processed |
|  |  |  | This question also did not perform well in face and content validity testing with experts. | foods within CFG-2019 healthy food choices guidance.<br>The updated question was tested in subsequent rounds of cognitive interviews in English and French and found to be easier to understand compared to previous iterations. |
| 3 | Lack of clarity about what to include or exclude/Food groupings | Sugary foods | The initial version of this question included example foods such as sugary breakfast cereals, cookies, and cakes (in French, des céréales sucrées pour le déjeuner, des biscuits, des gâteaux). In cognitive testing in French, some participants were uncertain whether the list of examples was exhaustive, or whether sugary snacks not specifically mentioned should be included. | To indicate that the list of examples was not exhaustive, “or other sugary foods” (“ou d’autres aliments sucrés”) was added after the list of examples.<br>This modification was tested in subsequent rounds of cognitive interviews in English and French and appeared to help participants to “think outside the box” and consider foods not specifically mentioned in the question. |
| 4 | Lack of clarity about what to include or exclude/<br>Foods consumed in multiple ways | Unsweetened milks<br>Oils | The screener question probing unsweetened milk consumption originally asked, “how often did you drink milk and unsweetened plant-based beverages”? (in French, “à quelle fréquence avez-vous bu du lait et des boissons d’origine végétale non | The phrases “drink” (“boire”) and “cook with or add” (“cuisiner ou ajouter”) were changed to the more general keyword, “consume” (“consommer”) to indicate that the foods could be consumed in multiple ways. Though using “consommer” appeared to resolve the |
|  |  |  | sucrées ”). In cognitive testing in both English and French, participants expressed uncertainty about whether to report only milk drunk in a glass or if it was appropriate to report milk used as an ingredient or component of a dish.<br>Similarly, the question probing oil consumption asked respondents, “how often did you cook with or add vegetable oils or soft margarines to your foods?” (in French, “combien de fois avez-vous cuisiné avec des margarines molles ou des huiles végétales[...] ou en avez-vous ajoutées à vos aliments?”). In cognitive testing in both English and French, several participants did not consider oils or margarines used outside of cooking, such as in salad dressing or spread on toast, until prompted by the interviewer. | issue in French, three participants in the second round of testing in English reported confusion with the word “consume”.<br>In the English version of the screener, the question was rephrased using the word “have” (e.g., “how often did you have milk and unsweetened plant-based beverages?”). A direct translation of the term “have” was not available in French, and the term “consommer” (to consume) did not elicit the same issues in the cognitive testing in French. Comprehensibility was prioritized over translational equivalency in this case, retaining “consommer” (“à quelle fréquence avez-vous consommé du lait de vache ou des boissons d’origine végétale non sucrées”) in French.<br>The modifications were tested in subsequent rounds of cognitive interviews in English and French, and the problems did not recur. |
| 5 | Keyword confusion | Unsweetened milks | In cognitive testing in French, several participants were uncertain whether “lait” (“milk”) referred only to cow’s milk or other types of milk, such as plant-based beverages or milk from other animals. This issue did not occur in cognitive testing in English. | To clarify that “lait” refers to cow’s milk, the phrase was replaced with “lait de vache” (“cow’s milk”) in the French version of the screener. To maintain translational equivalency, “milk” was replaced with “cow’s milk” in the English version of the screener.<br>The modifications were tested in the final rounds of cognitive interviews in English and French, and found to perform well.<br>This specification (inclusion of cow’s milk/lait de vache) was not included in the question on sweetened flavoured milks because the issue did not arise for that question, and so adding another keyword to the question was not warranted. |
| 6 | Keyword confusion | Animal-based proteins | The animal-based protein question originally queried consumption of “lean red meat” (in French, “viande rouge maigre”). In cognitive testing in French, several participants were uncertain which types of meats would be considered “lean” (“maigre”). This issue did not occur in cognitive testing in English. | The phrase “lean red meat” (“viande rouge maigre”) was replaced with the more specific keywords, “beef and pork” (“boeuf et porc”). Changes were made to both versions of the screener to maintain translational equivalency.<br>This wording was found to perform well in the final rounds of cognitive interviews in English and French. |
| 7 | Readability | Animal-based proteins | This question initially placed eggs at the end of a list of animal-based proteins including red meat, poultry, and shellfish. A participant in the first round of cognitive interviews in English explained they only skimmed the question after reading the first few examples and assumed only meat products should be included. They therefore did not consider eggs until prompted by the interviewer. | Since the other example foods listed were meat (and shellfish) products, eggs were moved to the front of the list of example food items.<br>This modification was tested in the second round of cognitive interviews in English and French, and the problem did not recur. |
| 8 | Readability | Salty snacks | Crackers were initially placed at the end of a list of salty snacks such as chips and pretzels. Several participants in the English interviews did not consider crackers in their responses, as they assumed the question was asking about foods conventionally viewed as “junk” foods. In contrast, crackers are generally considered “healthier”. | Crackers were moved to the front of the list of example food items.<br>This modification was tested in the final rounds of cognitive interviews in English and French, and the problem did not recur. |
| 9 | Readability | Plant-based proteins<br>Oils<br>Sweetened milks | Initial drafts of the screener included parentheses within some questions containing examples of specific foods that should be considered in responding. In cognitive interviews in French, several participants expressed that the parentheses made the question difficult to read and understand. They noted that they stopped reading when they encountered parentheses, causing foods listed within the parentheses to be overlooked. | In both the English and French versions, parentheses were removed or moved to the end of the question to enhance readability.<br>These modifications were tested in the second rounds of cognitive interviews in English and French and the problem did not recur. |
| 10 | Response errors | Vegetables<br>Whole grains | Two participants in the English interviews reported their responses as quantities rather than frequencies for the vegetable question, and one of those participants did the same for whole grains. Both referenced “servings” as specified by prior iterations of Canada’s Food Guide. | No changes were made to the screener, as respondents are instructed to report frequency in both the preamble and in each individual screener question. |

<sup>1</sup>The versions of the screener, in English and French, tested in the initial round of cognitive interviews, as well as in face and content validity testing with experts, are available in Supplementary File S2.

Many foods can be consumed in multiple ways. For example, milk can be consumed as a beverage or used in a sauce or added to a bowl of cereal. Similarly, oil can be used in cooking or as part of a salad dressing. In cognitive testing, some participants were unsure whether certain foods should be reported only if consumed by a particular method and may have overlooked other methods (Table 3, **example 4**). To address this ambiguity, general terms were used to encompass multiple methods of consumption (e.g., *have* instead of *drink* milk).

#### Keyword confusion

Some questions in the initial screener included keywords that were unclear or vague to participants, particularly in the testing in French, creating opportunities for misinterpretation (Table 3, **examples 5 and 6**). Lack of clarity was addressed by using more specific keywords (e.g., “lait de vache” instead of lait and “cow’ s milk” instead of milk) to describe the foods that should be included in each category.

### Readability

Readability issues occurred when the structure of a question hindered participants’ comprehension. When encountering lengthy lists of examples, participants tended to miss details and consequently provided inaccurate responses (Table 3, **examples 7 and 8**).

Including parentheses within a question to provide additional examples of particular food categories hindered readability, as participants felt the parentheses cued them to stop reading (Table 3, **example 9**). Such problems were alleviated through formatting changes and by re-wording and simplifying phrasing, for example, by reducing the number or changing the order of examples or breaking a single question into multiple questions.

#### Response option errors

There were a few cases in which participants reported quantity, rather than frequency of consumption, referencing serving sizes detailed by prior versions of CFG (Table 3, **example 10**). Because the screener instructs participants to report frequency in both the preamble and each question, no changes were made to address this issue.

### Face and content validity testing

For the testing in English, 21 content experts were invited and 13 accepted and for the testing in French, five content experts were invited and three accepted. These experts were academics, dietitians/nutritionists, and federal employees. Sociodemographic information was not collected from the content experts, but there was perceived variation in sex and race/ethnicity.

Overall, the content experts generally agreed that the initial screener questions reflected the guidance and were easy to understand. Issues raised were often consistent with those arising in the cognitive interviews. For example, the experts noted that the question assessing highly processed foods was overly complicated, with too many examples, such that it would not be understood by those with lower literacy levels. The experts’ feedback supported simplification of this question and others. Some experts suggested aligning the screener questions and structure more closely with the guidance, for instance, by ordering questions in a manner consistent with the guidance and including more detail, such as specifying additional example foods noted in the guidance.

### Refinements to the screener

Although questions were generally well-understood in both languages, changes were made to the order of the screener’ s questions as well as examples within each question to improve clarity and make the screener more intuitive for respondents (Table 3, **examples 7 and 8**). Rearranging questions and examples appeared to improve readability and comprehension by cueing respondents on which foods to include or exclude and emphasizing aspects of the question that were otherwise overlooked. In alignment with the guiding principle to develop a screener that is simple to use and the cognitive testing findings, changes were not made to address suggestions from the experts to order questions consistent with the guidance itself and to include more example foods.

More substantial modifications included the addition of a question to assess frequency of potato consumption (Table 3, **example 1**) because cognitive testing revealed that some participants tended not to include potatoes when asked about their vegetable consumption.

Instead, they viewed potatoes as a starchy food that might belong with grain foods. The question regarding highly processed foods was simplified to clarify what foods should and should not be included in responses.

In some cases, issues identified in one language entailed changes to the screener in both languages. For example, the expression “viande rouge maigre” (“lean red meat”) was unclear to participants in the cognitive testing in French, although the issue did not arise in English (Table 3, **example 6**). To ensure translational equivalency, “viande rouge maigre” (“lean red meat”) was replaced with “boeuf et porc” (“beef and pork”) in both versions of the screener. In other cases, changes were required in only one language. For example, in the cognitive testing in English, the term “have” (i.e., “how often did you have”) improved comprehension of the screener compared to terms like “drink”, which were overly specific and tended to limit respondents’ thought processes (Table 3, **example 4**). However, a direct translation of the term “have” was not available in French, and the term “consommer” (to consume) did not elicit the same issues in the French cognitive testing. Comprehensibility was prioritized over translational equivalency in this case, retaining “consommer” in French and “have” in English.

Formatting was used strategically, including line spacing and judicious use of bolding for emphasis; however, underlining and italics were avoided in the final screener to improve accessibility (City of Peterborough 2014; Kovac 2018).

### Final screener

The final version of the screener in each language includes 16 questions (Table 1). Nine assess consumption of healthy foods to “eat each day”, including fruit; vegetables; potatoes; animal-based protein foods; plant-based protein foods; yogurt, kefir, and cheese; unsweetened cow’ s milk and plant-based beverages; whole-grain foods; and margarine and vegetable oils. Seven questions assess “foods to limit”, including processed meat, fast food, sweetened cow’ s milk and plant-based beverages, other sugary beverages, sugary snacks, salty snacks, and refined grains. The final screener is available, in both English and French, in **Supplementary File S3**.

## Discussion

The Canadian Food Intake Screener/Questionnaire court canadien sur les apports alimentaires, available in English and French, rapidly assesses overall alignment of dietary intake with the healthy food choices recommendations in CFG-2019. The screener is intended for use in research with adults, aged 18-65 years, with marginal and higher health literacy. The screener was developed through an iterative process that included three rounds of cognitive interviews in each language along with ongoing feedback from advisors, as well as formal face and content validity testing with a separate panel of content experts. Results suggested that the screener was well understood in both languages, and informed refinements to question wording and screener structure to improve comprehension and minimize cognitive load. The screener can be completed in approximately five minutes.

Cognitive testing is a valuable method for identifying and correcting problems within a survey or screener and goes beyond conventional pre-testing to comprehensively examine respondents’ understanding of each item (Beatty and Willis 2007; Foddy 1996). Previous cognitive testing studies have revealed issues with ambiguous language and keyword misinterpretation (Bobrovitz et al, 2015; Eland et al, 2022), which was also observed in the present study. Small details, including individual words, can change a question’ s meaning; thus, the cognitive interviews were valuable to ensure the screener questions were understood as intended. Seemingly minor tweaks to the language and structure, such as changing “drink” to “have” (in the English version of the screener) and reordering examples, improved comprehension, helping to address the guiding principle related to an easy-to-use screener.

Cognitive interviews also exposed the challenge of designing questions that are specific enough to cue respondents on what to include and exclude, while not being so overly specific that they limit respondents’ thinking. Thompson et al. (2022) found a similar issue in the development of a food literacy questionnaire, wherein participants thought too narrowly about a particular context if the frame of reference was not well-defined. To address this issue in the current screener, example lists were kept as short and simple as possible, and in some cases, reference to “other” foods (e.g., other plant-based protein foods, other salty snacks) indicates that the list is not exhaustive. Providing exclusion criteria also appeared to help guide respondents on what should be included when responding to each question.

Potential uses of the screener are discussed in the accompanying paper (Hutchinson et al. 2022). The screener assesses intake over the past month, consistent with other screeners (Centers for Disease Control and Prevention n.d.; England et al. 2017; National Cancer Institute 2021; Wijnhoven et al. 2018). Adaptation to the past week or year is possible, but these time frames have not been evaluated. The screener captures the main elements of the healthy food choices recommendations from CFG-2019; however, given its brevity, it is not comprehensive. For example, it does not query all examples of highly processed foods (e.g., frozen entrées, sauces), though a range of foods noted as highly processed within the guidance are included (Health Canada 2022a). Given challenges in accurately measuring water consumption due to consumption throughout the day that is not structured around meals (Gandy 2015), the screener does not query frequency of water intake.

The screener was developed and evaluated for use with adults aged 18-65 years. It may be amenable to self-administration by older children without substantial modification.

However, recalling and reporting frequency of intake over the past month is likely to be cognitively challenging for younger children. It has been suggested that children can begin to conceptualize time at around ages seven to eight years and to self-report their own intake using a frequency-based instrument starting at around 10 years (Livingstone et al. 2004). Future research could evaluate the administration of the screener to caregivers as proxy reporters for younger children, similar to the implementation of 24-hour dietary recalls in national surveillance (Health Canada 2006; Health Canada 2017). Future research could also evaluate the screener for use with older adults. Evaluation of the screener used in the Canadian Longitudinal Study on Aging suggested differential performance among younger versus older adults (Gilsing et al. 2018), suggesting unique considerations related to age. Within that evaluation, considerations related to digital literacy among older adults were raised (Gilsing et al. 2018), but such concerns may be lessening in the digital era. Data from Statistics Canada indicate that the proportion of adults aged 65 years and older who accessed the Internet for personal use in the last three months increased from 48% in 2012 to 71% in 2018 (Statistics Canada 2019).

A guiding principle for the development and evaluation of the screener was to consider equivalence across population subgroups in capturing alignment with the healthy food choices recommendations. Equivalence relates to comparability (Boer et al. 2018; Frongillo et al., 2019; He and van de Vijver 2012) and can be threatened by construct bias, such that the construct intended to be measured is not the same across groups, as well as item bias, such that items have different meanings across groups (Boer et al. 2018; He and van de Vijver 2012). With respect to language, French-speaking advisors were involved throughout the process and versions in English and French were tested and modified simultaneously to maximize translational equivalency (Hebestreit et al. 2017; Kwon et al. 2020; Vieira et al. 2020). Nonetheless, English-speaking individuals were more heavily represented among the advisors and experts.

The screener queries a range of foods consumed by diverse populations. For example, the question on animal-based protein foods queries beef, pork, wild meat, chicken, and shellfish; and various types of grains, including rice, pasta, noodles, and breads, are listed. Further, reasonable variation in sociodemographic characteristics among cognitive testing participants was achieved, supporting relatively broad perspectives, including on the specific foods queried. However, the sample was skewed toward women, no participants identified as non-binary, and few identified as Black or Indigenous. The development process was informed by input from advisors who included researchers and practitioners from Canada, the USA, and Brazil, including experts in food-based dietary guidance and dietary assessment. Nonetheless, the field of nutrition and dietetics in Canada and elsewhere is relatively homogeneous (McBurney 2022) and structural barriers, including racism and heteronormativity, uphold this homogeneity (White 2018; Carter 2020; Burt et al. 2021; Joy and McSweeney-Flaherty 2022). Given the lack of representation of individuals with diverse and intersecting gender, racial/ethnic, and other identities, important perspectives related to dietary intake and its measurement among subgroups of the population may have been overlooked. Further evaluation of the screener with specific subgroups may thus be warranted to assess whether interpretation of the questions and their face and content validity are consistent. Moreover, it is critical to improve diversity and representation in the field to ensure that heterogeneity of the population is appropriately considered, as well as to heighten consideration of cross-context equivalence within dietary assessment.

Additional considerations are salient to the development of the screener. The sample size was consistent with recommendations for cognitive testing (Beatty and Willis 2007; Willis and Artino 2013; Willis and Miller 2011). However, some cognitive processes are difficult to verbalize, and it is possible that interviews may identify problems that would not occur when the screener is administered in the field or fail to identify issues that would emerge in the field (Beatty and Willis 2007). Informal coding has been used successfully in cognitive testing research, especially when time and resource constraints limit the ability to conduct full transcription of interview recordings (Willis 2005; Beatty and Willis 2007; Willis and Artino 2013;

Willis and Miller 2011). To assess the potential impact of more intensive coding, the recordings of the interviews in English were subsequently transcribed and coded using the Framework Method (Gale et al. 2017; Ritchie and Spencer 1994). Although additional instances of issues with comprehension of specific questions were noted, the issues themselves were identified by the informal coding, and no new issues likely to have prompted refinements to the screener were identified (Williams 2022).

## Conclusion

The Canadian Food Intake Screener allows for rapid assessment of the overall alignment of adults’ dietary intake with the healthy food choices recommendations within CFG-2019. Collaboration with a range of advisors, along with cognitive interviews and face and content validity testing, facilitated development of a simple screener in both English and French intended for use with adults with marginal and higher health literacy. The screener requires about five minutes to complete and is amenable to research and surveillance contexts in which it is not possible to conduct comprehensive dietary assessment. As is the case with the HEFI-2019 (Brassard et al. 2022a, 2022b), appropriate use of the screener can promote consistent assessment of alignment of adults’ dietary intakes with CFG-2019 healthy food choices recommendations. This is critical to creating an evidence base that can be synthesized to inform policies and programs to narrow the gap between current dietary intake and the guidance.

## Data Availability

Due to the nature of the data, they cannot be made openly available.

## Acknowledgements

The authors are grateful to the participants in the cognitive testing and the experts who participated in the face and content validity testing. The authors are also grateful to Kimberley J. Hernandez, Lisa-Anne Elvidge Munene, Sylvie St-Pierre, and other Health Canada employees who participated in and provided input on various aspects of the project; Dr. Hannah Tait Neufeld of the University of Waterloo who reviewed the screener for appropriateness for use with Indigenous populations and provided related advice; Dr. Gordon B. Willis of the United States National Cancer Institute for feedback on the cognitive interviewing protocol; and Sanaa Hussain of the University of Waterloo for assistance with referencing.

## Contributions

Conception and design: SIK, JMH, TEW, AMW, JH, AW, AM; analysis and interpretation of data: TEW, JMH, AMW, SIK, AB, CP, SL; drafting of the manuscript: JMH, TEW, SIK; revising the manuscript critically for important intellectual content: AMW, AB, CP, SL, KWD, BL, PMG, JH, AW, AM, MLCL, MJ, DLO, RP, JRS, JEV; final approval of the manuscript submitted: all authors. JMH and TEW contributed equally to this paper.

## Data availability

Due to the nature of the data, they cannot be made openly available.

