## Supplemental File S1 for "Development of the Canadian Food Intake Screener to assess alignment of adults’ dietary intake with the 2019 Canada’s Food Guide healthy food choices recommendations"

Hutchinson JM, Williams TE, et al.

**Supplementary File S1. Advisors who provided input on the development of the Canadian Food Intake Screener**

- Meghan Day, British Columbia Ministry of Health
- Kevin Dodd, U.S. National Cancer Institute
- Patricia Guenther, University of Utah
- Jess Haines, University of Guelph
- Mahsa Jessri, University of British Columbia
- Mary L'Abbé, University of Toronto
- Benoît Lamarche, Université Laval
- Simone Lemieux, Université Laval
- Maria Laura Louzada, University of São Paulo
- Dana Lee Olstad, University of Calgary
- Rachel Prowse, Memorial University of Newfoundland
- Janis Randall Simpson, University of Guelph
- Jill Reedy, U.S. National Cancer Institute
- Hassan Vatanparast, University of Saskatchewan
- Jennifer Vena, Alberta Health Services
