## Supplemental File S2 for "Development of the Canadian Food Intake Screener to assess alignment of adults’ dietary intake with the 2019 Canada’s Food Guide healthy food choices recommendations"

Hutchinson JM, Williams TE, et al.

**Supplementary File S2. Initial questions in the Canadian Food Intake Screener to assess alignment of adults' intake with the Canada Food Guide-2019 healthy food choices recommendations**

**NOTE: THIS IS THE VERSION OF THE SCREENER THAT WAS TESTED IN COGNITIVE INTERVIEWS AND FACE AND CONTENT VALIDITY TESTING.**

**SEE SUPPLEMENTARY FILE S3 FOR THE FINAL SCREENER.**

**NOTE: THIS DRAFT DOES NOT CONTAIN THE FRENCH VERSION OF THE SCREENER DUE TO SERVER LANGUAGE REQUIREMENTS. PLEASE CONTACT THE CORRESPONDING AUTHOR TO ACCESS THE FRENCH VERSION.**

DRAFT

These questions are about foods and beverages you ate or drank in the past month. When answering, please include meals and snacks consumed at home, at work or school, in restaurants, and anyplace else.

1. Over the past month, how often did you consume fresh, frozen, and canned fruit? *Do **not** include fruit juices or drinks*.
   - ☐ Never
   - ☐ 1 time in the past month
   - ☐ 2-3 times in the past month
   - ☐ 1-2 times per week
   - ☐ 3-4 times per week
   - ☐ 5-6 times per week
   - ☐ 1 time per day
   - ☐ 2-3 times per day
   - ☐ 4-5 times per day
   - ☐ 6 or more times per day

2. Over the past month, how often did you consume cooked, raw, frozen, and canned vegetables? *Do **not** include deep-fried vegetables or vegetable juices or drinks*.
   - ☐ Never
   - ☐ 1 time in the past month
   - ☐ 2-3 times in the past month
   - ☐ 1-2 times per week
   - ☐ 3-4 times per week

- 5-6 times per week
- 1 time per day
- 2-3 times per day
- 4-5 times per day
- 6 or more times per day

3. Over the <u>past month</u>, how often did you consume deep-fried foods and ready-to-heat or ready-to-eat dishes? Include frozen, canned, and packaged meat-based and vegetarian/vegan dishes, such as soups, pre-made pasta and rice dishes, and plant-based meats. *Do **not** include hot dogs, sausages, ham, corned beef, beef jerky or other deli or luncheon meats*.
   - Never
   - 1 time in the past month
   - 2-3 times in the past month
   - 1-2 times per week
   - 3-4 times per week
   - 5-6 times per week
   - 1 time per day
   - 2-3 times per day
   - 4-5 times per day
   - 6 or more times per day

DRAFT

*4.* Over the <u>past month</u>, how often did you consume hot dogs, sausages, ham, corned beef, beef jerky, and other deli or luncheon meats? *Do **not** include canned fish or canned poultry or packaged plant-based meats.*
   - Never
   - 1 time in the past month
   - 2-3 times in the past month
   - 1-2 times per week
   - 3-4 times per week
   - 5-6 times per week
   - 1 time per day
   - 2-3 times per day
   - 4-5 times per day
   - 6 or more times per day

5. Over the <u>past month</u>, how often did you consume lean red meat or pork, wild game, poultry, fish, shellfish, and eggs? Include canned fish and canned poultry. *Do **not** include hot dogs, sausages, ham, corned beef, beef jerky, or other deli or luncheon meats*.
   - Never
   - 1 time in the past month
   - 2-3 times in the past month

☐ 1-2 times per week
☐ 3-4 times per week
☐ 5-6 times per week
☐ 1 time per day
☐ 2-3 times per day
☐ 4-5 times per day
☐ 6 or more times per day

6. Over the past month, how often did you consume nuts (including nut butters), seeds, tofu, beans (e.g., chickpeas, hummus, lentils, black beans), and other plant-based sources of protein? *Do **not** include green beans or ready-to-heat foods, such as pre-made veggie burgers and packaged plant-based meats*.

☐ Never
☐ 1 time in the past month
☐ 2-3 times in the past month
☐ 1-2 times per week
☐ 3-4 times per week
☐ 5-6 times per week
☐ 1 time per day
☐ 2-3 times per day
☐ 4-5 times per day
☐ 6 or more times per day

DRAFT

7. Over the past month, how often did you drink milk and unsweetened plant-based beverages (e.g., soy, almond, and oat milk)? *Do **not** include small amounts in coffee or tea, or chocolate and other flavoured milk*.

☐ Never
☐ 1 time in the past month
☐ 2-3 times in the past month
☐ 1-2 times per week
☐ 3-4 times per week
☐ 5-6 times per week
☐ 1 time per day
☐ 2-3 times per day
☐ 4-5 times per day
☐ 6 or more times per day

*8.* Over the past month, how often did you consume yogurt, kefir, and cheese?

☐ Never
☐ 1 time in the past month
☐ 2-3 times in the past month
☐ 1-2 times per week
☐ 3-4 times per week

- ☐ 5-6 times per week
- ☐ 1 time per day
- ☐ 2-3 times per day
- ☐ 4-5 times per day
- ☐ 6 or more times per day

9. Over the past month, how often did you drink flavoured milk, sweetened plant-based beverages (e.g., soy, almond, and oat milk), and sweetened coffee or tea (including bottled)? *Do **not** include diet or sugar-free beverages*.
   - ☐ Never
   - ☐ 1 time in the past month
   - ☐ 2-3 times in the past month
   - ☐ 1-2 times per week
   - ☐ 3-4 times per week
   - ☐ 5-6 times per week
   - ☐ 1 time per day
   - ☐ 2-3 times per day
   - ☐ 4-5 times per day
   - ☐ 6 or more times per day

DRAFT

10. Over the past month, how often did you drink soda or pop containing sugar, fruit juice and fruit-flavoured drinks, sweetened sports drinks, and sweetened waters? *Do **not** include diet or sugar-free beverages, such as diet soda and plain water*.
   - ☐ Never
   - ☐ 1 time in the past month
   - ☐ 2-3 times in the past month
   - ☐ 1-2 times per week
   - ☐ 3-4 times per week
   - ☐ 5-6 times per week
   - ☐ 1 time per day
   - ☐ 2-3 times per day
   - ☐ 4-5 times per day
   - ☐ 6 or more times per day

11. Over the past month, how often did you consume sugary breakfast cereals, cookies, cakes, pastries, granola bars, ice cream, candy, and chocolate?
   - ☐ Never
   - ☐ 1 time in the past month
   - ☐ 2-3 times in the past month
   - ☐ 1-2 times per week
   - ☐ 3-4 times per week
   - ☐ 5-6 times per week
   - ☐ 1 time per day

- ☐ 2-3 times per day
- ☐ 4-5 times per day
- ☐ 6 or more times per day

12. Over the past month, how often did you consume chips, pretzels, popcorn, crackers, and other salty snacks?
    - ☐ Never
    - ☐ 1 time in the past month
    - ☐ 2-3 times in the past month
    - ☐ 1-2 times per week
    - ☐ 3-4 times per week
    - ☐ 5-6 times per week
    - ☐ 1 time per day
    - ☐ 2-3 times per day
    - ☐ 4-5 times per day
    - ☐ 6 or more times per day

13. Over the past month, how often did you consume white breads, bagels, rice, pasta, and noodles? *Do **not** include whole wheat or whole grain foods*.
    - ☐ Never
    - ☐ 1 time in the past month
    - ☐ 2-3 times in the past month
    - ☐ 1-2 times per week
    - ☐ 3-4 times per week
    - ☐ 5-6 times per week
    - ☐ 1 time per day
    - ☐ 2-3 times per day
    - ☐ 4-5 times per day
    - ☐ 6 or more times per day

DRAFT

14. Over the past month, how often did you consume whole wheat or whole grain breads, rice, pasta, noodles, and cereals? *Do **not** include white breads, rice, pasta, noodles, or cereals*.
    - ☐ Never
    - ☐ 1 time in the past month
    - ☐ 2-3 times in the past month
    - ☐ 1-2 times per week
    - ☐ 3-4 times per week
    - ☐ 5-6 times per week
    - ☐ 1 time per day
    - ☐ 2-3 times per day
    - ☐ 4-5 times per day

- ☐ 6 or more times per day

*15.* Over the past month, how often did you cook with or add vegetable oils (e.g., canola, olive, sunflower) or soft margarines to your foods? *Do **not** include coconut oil, palm oil, or butter*.

- ☐ Never
- ☐ 1 time in the past month
- ☐ 2-3 times in the past month
- ☐ 1-2 times per week
- ☐ 3-4 times per week
- ☐ 5-6 times per week
- ☐ 1 time per day
- ☐ 2-3 times per day
- ☐ 4-5 times per day
- ☐ 6 or more times per day

DRAFT
