## Supplemental File S3 for "Development of the Canadian Food Intake Screener to assess alignment of adults’ dietary intake with the 2019 Canada’s Food Guide healthy food choices recommendations"

Hutchinson JM, Williams TE, et al.

**Supplementary File S3. Canadian Food Intake Screener**

**NOTE: THIS SCREENER DOES NOT CONTAIN THE FRENCH VERSION DUE TO SERVER LANGUAGE REQUIREMENTS. PLEASE CONTACT THE CORRESPONDING AUTHOR TO ACCESS THE FRENCH VERSION.**

**Canadian Food Intake Screener**

These questions are about foods and beverages you ate or drank in the past month, that is, the past 30 days. When answering, please include meals and snacks consumed at home, at work or school, in restaurants, and anyplace else.

1. Over the past month, how often did you eat fresh, frozen, canned, or dried fruit?

   **Do not include** fruit juices and drinks.

   - ❑ Never
   - ❑ 1 time in the past month
   - ❑ 2-3 times in the past month
   - ❑ 1-2 times per week
   - ❑ 3-4 times per week
   - ❑ 5-6 times per week
   - ❑ 1 time per day
   - ❑ 2-3 times per day
   - ❑ 4-5 times per day
   - ❑ 6 or more times per day

2. Over the past month, how often did you eat potatoes, including baked, boiled, or mashed potatoes, or sweet potatoes?

   **Do not include** french fries, poutine, home fries, or hash browns.

   - ❑ Never
   - ❑ 1 time in the past month
   - ❑ 2-3 times in the past month
   - ❑ 1-2 times per week
   - ❑ 3-4 times per week
   - ❑ 5-6 times per week
   - ❑ 1 time per day
   - ❑ 2-3 times per day
   - ❑ 4-5 times per day
   - ❑ 6 or more times per day

3. Over the past month, how often did you eat fresh, cooked, frozen, or canned vegetables?

   **Do not include** potatoes, french fries, poutine, or other deep-fried vegetables, or vegetable juices and drinks.

   - ❏ Never
   - ❏ 1 time in the past month
   - ❏ 2-3 times in the past month
   - ❏ 1-2 times per week
   - ❏ 3-4 times per week
   - ❏ 5-6 times per week
   - ❏ 1 time per day
   - ❏ 2-3 times per day
   - ❏ 4-5 times per day
   - ❏ 6 or more times per day

4. Over the past month, how often did you eat food from fast food restaurants, such as burgers, french fries, poutine, pizza, submarine sandwiches, fried chicken, burritos, or tacos?

   - ❏ Never
   - ❏ 1 time in the past month
   - ❏ 2-3 times in the past month
   - ❏ 1-2 times per week
   - ❏ 3-4 times per week
   - ❏ 5-6 times per week
   - ❏ 1 time per day
   - ❏ 2-3 times per day
   - ❏ 4-5 times per day
   - ❏ 6 or more times per day

5. Over the past month, how often did you eat hot dogs, sausages, beef jerky, bacon, ham or other deli or luncheon meats?

   **Do not include** fast food, canned fish, canned poultry, or packaged veggie burgers and plant-based meats.

   - ❑ Never
   - ❑ 1 time in the past month
   - ❑ 2-3 times in the past month
   - ❑ 1-2 times per week
   - ❑ 3-4 times per week
   - ❑ 5-6 times per week
   - ❑ 1 time per day
   - ❑ 2-3 times per day
   - ❑ 4-5 times per day
   - ❑ 6 or more times per day

6. Over the past month, how often did you eat eggs, beef, pork, wild meat, chicken or other poultry, fish, shellfish, or other animal-based sources of protein? Include canned fish and canned poultry.

   **Do not include** fast food, hot dogs, sausages, beef jerky, bacon, ham, or other deli or luncheon meats.

   - ❑ Never
   - ❑ 1 time in the past month
   - ❑ 2-3 times in the past month
   - ❑ 1-2 times per week
   - ❑ 3-4 times per week
   - ❑ 5-6 times per week
   - ❑ 1 time per day
   - ❑ 2-3 times per day
   - ❑ 4-5 times per day
   - ❑ 6 or more times per day

7. Over the past month, how often did you eat nuts, seeds, tofu, beans and lentils, peanut butter or other nut butters, or other plant-based sources of protein?

   **Do not include** green beans or packaged veggie burgers and plant-based meats.

   - ❑ Never
   - ❑ 1 time in the past month
   - ❑ 2-3 times in the past month
   - ❑ 1-2 times per week
   - ❑ 3-4 times per week
   - ❑ 5-6 times per week
   - ❑ 1 time per day
   - ❑ 2-3 times per day
   - ❑ 4-5 times per day
   - ❑ 6 or more times per day

8. Over the past month, how often did you eat yogurt, kefir, or cheese?

   - ❑ Never
   - ❑ 1 time in the past month
   - ❑ 2-3 times in the past month
   - ❑ 1-2 times per week
   - ❑ 3-4 times per week
   - ❑ 5-6 times per week
   - ❑ 1 time per day
   - ❑ 2-3 times per day
   - ❑ 4-5 times per day
   - ❑ 6 or more times per day

9. Over the past month, how often did you have **white** cows' milk or **unsweetened** plant-based beverages (e.g., soy, almond, or oat milk)?

   **Do not include** small amounts in coffee or tea, or chocolate and other sweetened milk.

   - ❑ Never
   - ❑ 1 time in the past month
   - ❑ 2-3 times in the past month
   - ❑ 1-2 times per week
   - ❑ 3-4 times per week
   - ❑ 5-6 times per week
   - ❑ 1 time per day
   - ❑ 2-3 times per day
   - ❑ 4-5 times per day
   - ❑ 6 or more times per day

10. Over the past month, how often did you have chocolate milk or other **flavoured** milk or **sweetened** plant-based beverages (e.g., soy, almond, or oat milk)?

**Do not include** small amounts in coffee or tea, or diet/artificially sweetened or sugar-free beverages.

- ❑ Never
- ❑ 1 time in the past month
- ❑ 2-3 times in the past month
- ❑ 1-2 times per week
- ❑ 3-4 times per week
- ❑ 5-6 times per week
- ❑ 1 time per day
- ❑ 2-3 times per day
- ❑ 4-5 times per day
- ❑ 6 or more times per day

11. Over the past month, how often did you drink fruit juice, fruit-flavoured drinks, soda or pop, **sweetened** sports or energy drinks, **sweetened** hot or iced coffee or tea, or **sweetened** waters?

**Do not include** diet/artificially sweetened or sugar-free beverages, such as diet soda.

- ❑ Never
- ❑ 1 time in the past month
- ❑ 2-3 times in the past month
- ❑ 1-2 times per week
- ❑ 3-4 times per week
- ❑ 5-6 times per week
- ❑ 1 time per day
- ❑ 2-3 times per day
- ❑ 4-5 times per day
- ❑ 6 or more times per day

12. Over the past month, how often did you eat cookies, cakes, muffins, pastries, granola bars, protein bars, ice cream, candy, chocolate, sugary breakfast cereals, or other sugary foods?

- ❑ Never
- ❑ 1 time in the past month
- ❑ 2-3 times in the past month
- ❑ 1-2 times per week
- ❑ 3-4 times per week
- ❑ 5-6 times per week
- ❑ 1 time per day
- ❑ 2-3 times per day
- ❑ 4-5 times per day
- ❑ 6 or more times per day

13. Over the past month, how often did you eat crackers, chips, pretzels, popcorn, or other salty snacks?

- ❑ Never
- ❑ 1 time in the past month
- ❑ 2-3 times in the past month
- ❑ 1-2 times per week
- ❑ 3-4 times per week
- ❑ 5-6 times per week
- ❑ 1 time per day
- ❑ 2-3 times per day
- ❑ 4-5 times per day
- ❑ 6 or more times per day

14. Over the past month, how often did you eat **white** breads, bagels, rice, pasta, noodles, or other refined grains, such as breakfast cereals?

**Do not include** whole wheat or whole grain foods.

- ❑ Never
- ❑ 1 time in the past month
- ❑ 2-3 times in the past month
- ❑ 1-2 times per week
- ❑ 3-4 times per week
- ❑ 5-6 times per week
- ❑ 1 time per day
- ❑ 2-3 times per day
- ❑ 4-5 times per day
- ❑ 6 or more times per day

15. Over the past month, how often did you eat **whole wheat or whole grain** breads, bagels, pasta, noodles, quinoa, oats, brown or wild rice, breakfast cereals, or other whole wheat or whole grain foods?

    **Do not include** white breads, bagels, pasta, noodles, rice, or refined breakfast cereals.

    - ❑ Never
    - ❑ 1 time in the past month
    - ❑ 2-3 times in the past month
    - ❑ 1-2 times per week
    - ❑ 3-4 times per week
    - ❑ 5-6 times per week
    - ❑ 1 time per day
    - ❑ 2-3 times per day
    - ❑ 4-5 times per day
    - ❑ 6 or more times per day

16. Over the past month, how often did you have margarine or vegetable oils (e.g., olive, canola, or sunflower oil)?

    Do **not** include lard, coconut oil, palm oil, or butter.

    - ❑ Never
    - ❑ 1 time in the past month
    - ❑ 2-3 times in the past month
    - ❑ 1-2 times per week
    - ❑ 3-4 times per week
    - ❑ 5-6 times per week
    - ❑ 1 time per day
    - ❑ 2-3 times per day
    - ❑ 4-5 times per day
    - ❑ 6 or more times per day
